# Clonal Hematopoiesis of Indeterminate Potential in Patients with Chronic Thromboembolic Pulmonary Hypertension

**DOI:** 10.1101/2023.05.16.23290071

**Authors:** Chao Liu, Chun-Yan Cheng, Tian-Yu Lian, Yu-Ping Zhou, Yin-Jian Yang, Si-Jin Zhang, Yun-Peng Wei, Yong-Jian Zhu, Lu-Hong Qiu, Bao-Chen Qiu, Li-Yan Ren, Jing-Si Ma, Ruo-Nan Li, Jia Wang, Ying-Hui-Zi Shen, Zhi-Yan Han, Jing-Hui Li, Qin-Hua Zhao, Lan Wang, Xi-Qi Xu, Kai Sun, Duo-Lao Wang, Ze-Jian Zhang, Zhi-Cheng Jing

**Affiliations:** Department of Cardiology, State Key Laboratory of Complex Severe and Rare Diseases, Peking Union Medical College Hospital, Chinese Academy of Medical Sciences and Peking Union Medical College, Beijing, China; Medical Research Center, State Key Laboratory of Complex Severe and Rare Diseases, Peking Union Medical College Hospital, Chinese Academy of Medical Sciences and Peking Union Medical College, Beijing, China; Department of Cardiovascular Medicine, Ruijin Hospital Affiliated to Shanghai Jiao Tong University School of Medicine, Shanghai, China; Department of Cardiology, The First Affiliated Hospital of Zhengzhou University, Zhengzhou, China; School of Pharmacy, Henan University, Henan, China; Department of Medical Laboratory, Weifang Medical University, Weifang, Shandong, China; Department of Cardiovascular Medicine, The First Hospital of Jilin University, Changchun, China; State Key Laboratory of Cardiovascular Disease and FuWai Hospital, Chinese Academy of Medical Sciences and Peking Union Medical College, Beijing, China; Department of Cardio-Pulmonary Circulation, Shanghai Pulmonary Hospital, Tongji University, Shanghai, China; Department of Clinical Sciences, Liverpool School of Tropical Medicine, Liverpool, United Kingdom

**Author notes:** These authors contributed equally to this work as the co-first authors (Chao Liu, Chun-Yan Cheng, Tian-Yu Lian). Drs Zhi-Cheng Jing and Ze-Jian Zhang contributed equally to this study as the co-corresponding authors., **Address for Correspondence**: Zhi-Cheng Jing, MD, PhD, Department of Cardiology, State Key Laboratory of Complex Severe and Rare Diseases, Peking Union Medical College Hospital, Chinese Academy of Medical Sciences and Peking Union Medical College, No. 1 Shuaifuyuan Wangfujing, Dongcheng District, Beijing 100730, China., Ze-Jian Zhang, PhD, Medical Research Center, State Key Laboratory of Complex Severe and Rare, Diseases, Peking Union Medical College Hospital, Chinese Academy of Medical Sciences and Peking Union Medical College, No. 1 Shuaifuyuan Wangfujing, Dongcheng District, Beijing 100730, China.

**Keywords:** CHIP, CTEPH, inflammation, clinical outcome, high-confidence CHIP calls strategies

## Abstract

**Background:** The pathogenesis of chronic thromboembolic pulmonary hypertension (CTEPH) is complex and multifactorial, with growing evidence indicating the involvement of hematologic disorders. Clonal hematopoiesis of indeterminate potential (CHIP) has recently been associated with increased risks of both hematologic malignancies and cardiovascular diseases. The CHIP in patients with CTEPH and its clinical relevance remain undetermined.

**Methods:** We performed a step-wise calling method on the next-generation sequencing data from 499 CTEPH patients referred to three centers between October 2006 and December 2021 to identify CHIP mutations. We associated CHIP with all-cause mortality in patients with CTEPH. To provide potential mechanistic insights, the associations between CHIP and inflammation characteristics reflected by circulating cytokines and IgG galactosylation, a hallmark of inflammatory state in diseases were also determined.

**Results:** Total 51 (10.2%) patients with CTEPH carried at least one CHIP mutation at a variant allele frequency of ≥ 2%, and the most common mutations were among *DNMT3A*, *RUN1* and *STAG2*. During a mean follow-up time of 55 months, deaths occurred in 21 patients (42.9%) in the CHIP group and 105 patients (24.3%) in the non-CHIP group, contributing to the 5-year survival rate of 65.3% in the CHIP group and 81.9% in the non-CHIP group (*P* < 0.001 for log-rank test). The association of CHIP with mortality remained robust in the fully adjusted model (HR: 3.447; 95% CI: 1.747 – 6.803; *P* < 0.001). Besides, patients in the CHIP group showed higher circulating IL-1beta and IL-6 and lower IL-4 and IgG galactosylation compared with the non-CHIP group.

**Conclusions:** CHIP is enriched in CTEPH patients and is associated with a worse prognosis in CTEPH. Mechanically, patients in the CHIP group showed a more severe inflammatory state.

**Clinical Perspective:** *What is new?:*

- Clonal hematopoiesis of indeterminate potential (CHIP) mutations are enriched in chronic thromboembolic pulmonary hypertension (CTEPH) patients, with most common mutated genes being *DNMT3A*, *RUN1* and *STAG2*.
- CHIP is associated with worse clinical outcomes in CTEPH.
- CHIP is associated with more severe inflammation state mediated by myeloid as well as lymphoid cells in patients with CTEPH.

*What are the clinical implications?:*

- CHIP might be a risk factor for CTEPH, suggesting a relationship between CTEPH and hematopoietic disorders.
- CHIP represented an additional disease component in CTEPH that independently impacts prognosis.
- CHIP might be a potential target for personalized medicine and an indicator of benefit from anti-inflammatory therapies for CTEPH patients.

## Introduction

Chronic thromboembolic pulmonary hypertension (CTEPH) is a long-term and life-threatening complication of pulmonary embolism, characterized by chronic obstruction of pulmonary vasculature with organized and fibrotic thrombi and vascular remodeling^1, 2^. Increasing evidence showed that a variety of factors including inflammation^3–6^, coagulation dysregulation^7, 8^, genetics^9, 10^, *etc.* played important roles in the development of CTEPH, suggesting the complexity and multifactorial nature of pathogenesis involved.

Recently, there is more and more evidence supporting the involvement of hematologic disorders especially myeloproliferative neoplasms (MPN) in the pathogenesis of CTEPH, including higher rates of thrombotic events as well as recurrent venous thromboembolism in patients with MPN^11, 12^, the elevated prevalence of CTEPH in MPN patients^13^, and increased mortality after pulmonary endarterectomy (PEA) in CTEPH patients complicated with MPN^14^. Clonal hematopoiesis of indeterminate potential (CHIP), defined as the presence of an expanded somatic blood cell clone in persons without other hematologic diseases, is an early stage of MPN and hematologic cancers^15, 16^. The presence of CHIP was shown to increase with age and is significantly associated with the higher prevalence and worse prognosis of cardiovascular diseases^17–21^. However, the relationship between CHIP and CTEPH is yet to be investigated.

The present study aimed to assess the prevalence of CHIP-driver mutations in patients with CTEPH and, more importantly, to associate its presence with clinical outcomes.

## Method

### Study Population

From October 2006 to December 2021, all consecutive patients with definite diagnosis of CTEPH in three national pulmonary hypertension (PH) referral centers: Shanghai Pulmonary Hospital with patients mainly from south China and FuWai Hospital and Peking Union Medical College Hospital in Beijing with patients mainly from north China, were included in this study. For the diagnosis of CTEPH^22^, three criteria had to be satisfied: (1) at least three months of effective anticoagulation, including warfarin or new oral anticoagulant drugs (rivaroxaban or dabigatran); (2) typical imaging characteristics of CTEPH assessed by computed tomography pulmonary angiography and/or direct pulmonary angiography; and (3) confirmed pre-capillary PH defined as mean pulmonary artery pressure of ≥ 25 mm Hg and pulmonary arterial wedge pressure of ≤ 15 mm Hg. The hemodynamic evaluation was performed by right heart catheterization. Patients without available blood samples and therefore lacking sequencing data and CHIP information were excluded. According to the definition of CHIP, patients with previous hematologic cancers or other hematologic diseases were also excluded. The ethics review boards of the Shanghai Pulmonary Hospital, FuWai Hospital and Peking Union Medical College Hospital approved the protocols. All patients provided written informed consent. The study complies with the Declaration of Helsinki.

### Next-Generation Sequencing

Genomic DNA samples were either whole-exome sequencing (WES) or whole-genome sequencing (WGS) according to standard protocols. The WGS was performed at WuXi Apptech (Shanghai, China) using a PCR-free library preparation and paired-end (2 × 150 bp) sequencing on the HiSeq X Ten platform (Illumina, Inc., San Diego, CA, USA) to yield a minimum of 35 × coverage. The WES was performed at Novogene Corporation (Beijing, China) using the Agilent SureSelect Human All Exon kit (50 MB or 60 MB) and sequencing on the Illumina HiSeq instruments to provide a mean coverage of more than 100 × on the target regions of every sample. Prior to sequencing, samples were randomized to minimize batch effects. We used two sequencing platforms (WES and WGS) to genotype the CTEPH cases. To reduce systematic errors and alignment artifacts of the two sequencing platform, we restricted our call for variants to RefSeq coding sequences and called WGS together with WES using very strict criteria detailed below.

### Variant Calling and Annotation Strategies

We used previously established methods to identify CHIP, which were described in detail elsewhere^23^. In the first step, Mutect2, Genome Analysis Toolkit was used as a somatic variant calling pipeline to scan aligned sequencing files for putative somatic variants^24^. Briefly, Mutect2 was run in ‘tumor-only’ mode with default parameters, over a list of variants within 74 driver genes previously associated with CHIP^19^ (Table S1), as well as those new variants reported as somatic mutations in hematologic cancers at least seven times in COSMIC. Only non-synonymous exonic or splicing variants were considered. We removed variants with a total read depth of less than 15, variants with minimum read depth for the alternate allele of less than three, and variants lacking variant support in both forward and reverse sequencing reads. We also removed variants below the 2% variant allele frequency (VAF) threshold conventionally used to define CHIP. Variants occurring in 5% of all studied patients were considered probable recurrent sequencing artifacts. For this, the association of each variant group with age, an established strong correlate of CHIP was tested. Variants that were not associated with age at even a suggestive significance of *P* < 0.10 were removed from the dataset as they were suspected to represent sequencing artifacts^23^. To filter out potential germline variants, we conducted a binomial test across all variants to determine whether the measured read depth for the variant is statistically different from half of the sum of all sequencing reads at that site, as would be true for heterozygous germline variants. Variants that failed the binomial test at *P* < 0.01 would be removed from the dataset.

### Data Collection and Follow-up

Data on patient characteristics at first admission for CTEPH diagnosis were collected, including demographic information, clinical symptoms, medical history, World Health Organization functional class (WHO FC), 6-minute walking distance (6MWD), hematological testing, invasive hemodynamics, and treatments. Blood samples for genetic sequencing and circulating inflammation characteristics testing were collected during the first diagnostic right heart catheterization.

Patients were followed up via clinic visits or by phone or internet interviews, and the observation period was from the initial diagnosis of CTEPH to January 2022 or the latest visit. The single clinical endpoint was all-cause mortality.

### Inflammatory Characteristics Measurement

The levels of four circulating inflammatory markers were determined in the plasma of CTEPH patients using commercially available bead-based multiplex Luminex assays employing the Luminex 200 system for acquisition. Standard curves were included in the kits. The procedure was according to the manufacturer’s protocol and previous literature^25^.

The inflammatory marker of IgG Gal-ratio (distribution of IgG galactosylation) profiling is performed according to published protocols^26, 27^. Briefly, IgG was isolated from plasma samples using 96-well Protein A Spin Plates, and the glycans on IgG were enzymatically released by PNGase F. The released IgG N-glycans were then purified by porous graphitized carbon and analyzed by matrix-assisted laser desorption/ionization time of flight mass spectrometry (MALDI-TOF-MS) (Bruker Daltonics, Bremen). The processing of MALDI-TOF-MS raw data and IgG N-glycan identification was according to our previous work^26, 27^. The inflammatory marker of IgG Gal-ratio was obtained by calculating the relative intensities of agalactosylated (H3N4F1, G0) vs. monogalactosyl (H4N4F1, G1) and digalactosyl (H5N4F1, G2) N-glycans according to the formula of Gal-ratio = G0/(G1 +2 × G2) as previously reported^26, 27^.

### Statistical Analysis

Adherence to a Gaussian distribution was determined using the Kolmogorov-Smirnov test. Normally distributed data were described as mean SD, and the independent samples Student’s t-test was used to compare continuous variables between two groups. In the case of skewed distribution, data were described as median (IQR) and the nonparametric Mann-Whitney U-test was used for group comparisons. Qualitative variables were described with n (%) and analyzed using the Fisher exact test. Multivariable Cox proportional-hazards regression models were used to calculate hazard ratios (HR) for event rates, whereas Kaplan-Meier event rates and log-rank tests were used to compare survival estimates. All reported tests were 2-tailed, and the threshold for statistical significance was 0.05. The analyses were performed with SPSS 26 and GraphPad Prism 8.

## Results

### Study Population and baseline characteristics

The screening and inclusion of patients with CTEPH are presented in Figure 1. Initially, 567 patients suspected of having CTEPH were screened. Among them, eight patients were excluded due to having other aetiologies of pulmonary artery stenosis (four with pulmonary arteritis, two with congenital pulmonary artery stenosis and two with pulmonary artery sarcomas). Fifty-four patients without available blood samples and six patients with previous hematologic diseases were further excluded. Finally, 499 patients were included in the analysis.

**Figure 1.**
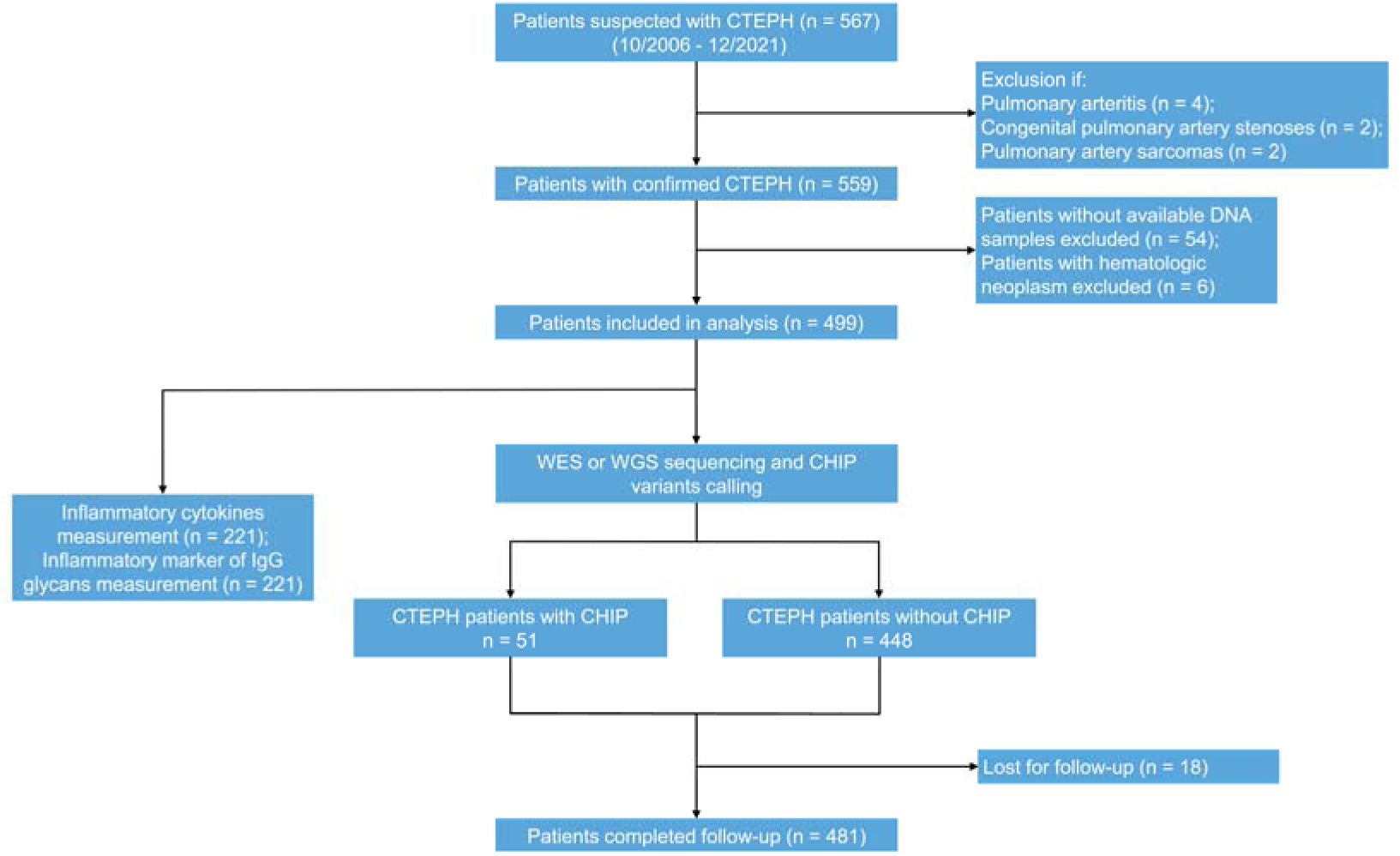
Diagram showing the selection of patients with CTEPH in the present study. CHIP = clonal hematopoiesis of indeterminate potential; CTEPH = chronic thromboembolic pulmonary hypertension. WES: whole-exome sequencing; WGS: whole-genome sequencing.

Baseline characteristics are provided in Table 1: 248 patients (49.7%) were male; mean patient age was 54.4 ± 14.8 years; 468 patients (93.8%) received PH-targeted drugs therapy; 273 patients (54.7%) underwent PEA or BPA treatment. Specifically, 198 patients (39.7%) underwent BPA, 106 patients (21.2%) underwent PEA and 31 patients (6.2%) received BPA and PEA bridging therapy.

**Table 1.** Characteristics of CTEPH patients stratified according to CHIP status.

|  | **Total cohort**<br>**(n = 499)** | **CHIP**<br>**(n = 51)** | **Non-CHIP**<br>**(n = 448)** | **P**<br>**value** |
| --- | --- | --- | --- | --- |
| **Demographic variables** |  |  |  |  |
| Age, y | 54.4 ± 14.8 | 58.4 ± 13.4 | 54.0 ± 14.9 | **0.045** |
| Male | 248 (49.7) | 23 (44.9) | 225 (50.2) | 0.488 |
| Diagnostic delay, mo | 24.6 (11.8 - 60.9) | 26.2 (11.6 - 61.7) | 24.5 (11.6 - 60.9) | 0.962 |
| Blood group non-O | 395 (79.2) | 42 (81.8) | 353 (78.8) | 0.553 |
| Prior VTE history | 360 (72.1) | 39 (76.5) | 321 (71.7) | 0.467 |
| Recurrent VTE history | 34 (6.8) | 4 (7.8) | 30 (6.7) | 0.758 |
| **Comorbid conditions** |  |  |  |  |
| Thrombophilia disorders | 38 (7.6) | 3 (5.9) | 35 (7.8) | 0.622 |
| APS | 30 (6.0) | 3 (5.9) | 27 (6.0) | 0.967 |
| Pacemaker | 1 (0.2) | 0 (0.0) | 1 (0.2) | 0.736 |
| Splenectomy | 4 (0.8) | 0 (0.0) | 4 (0.9) | 0.498 |
| Active cancer | 6 (1.2) | 0 (0.0) | 6 (1.3) | 0.406 |
| Renal insufficiency | 23 (4.6) | 4 (7.8) | 19 (4.2) | 0.245 |
| Coronary artery disease | 29 (5.8) | 4 (7.8) | 25 (5.6) | 0.513 |
| Ischemic stroke | 27 (5.4) | 1 (2.0) | 26 (5.8) | 0.25 |
| Diabetes | 23 (4.6) | 2 (3.9) | 21 (4.7) | 0.805 |
| Anemia | 26 (5.2) | 3 (5.9) | 23 (5.1) | 0.82 |
| **Clinical variables** |  |  |  |  |
| WHO FC (III - IV) | 320 (64.1) | 42 (82.4) | 278 (62.1) | **0.004** |
| 6MWD, m | 384.2 ± 116.3 | 335.7 ± 127.0 | 389.3 ± 114.2 | **0.013** |
| NT-proBNP, pg/mL | 1,161.0 (292.0 - 2,863.3) | 1,886.0 (754.8 - 3,787.9) | 1,104.5 (268.0 - 2,820.0) | **0.019** |
| D-dimer, ng/mL | 450.0 (200.0 - 1020.0) | 610.0 (330.0 - 1540.0) | 420.0 (190.0 - 970.0) | **0.012** |
| Hgb, mg/L | 146.8 ± 21.5 | 143.1 ± 23.1 | 147.2 ± 21.3 | 0.348 |
| Plt, /μL | 198.9 ± 70.3 | 194.1 ± 72.6 | 199.5 ± 70.1 | 0.663 |
| WBC, /mL | 6.3 ± 2.0 | 6.4 ± 2.2 | 6.3 ± 2.0 | 0.691 |
| **Hemodynamics** |  |  |  |  |
| RAP, mm Hg | 8.3 ± 5.4 | 9.7 ± 6.9 | 8.1 ± 5.2 | 0.142 |
| mPAP, mm Hg | 49.1 ± 12.1 | 48.8 ± 10.9 | 49.2 ± 12.3 | 0.839 |
| PAWP, mm Hg | 9.8 ± 3.4 | 10.3 ± 4.0 | 9.7 ± 3.3 | 0.324 |
| PVR, Wood units | 9.5 ± 4.7 | 10.1 ± 4.3 | 9.5 ± 4.7 | 0.382 |
| CI, L/min/m<sup>2</sup> | 2.6 ± 0.7 | 2.3 ± 0.5 | 2.6 ± 0.7 | **0.015** |
| SvO<sub>2</sub>, % | 62.1 ± 8.5 | 58.4 ± 10.0 | 62.5 ± 8.3 | **0.002** |
| Treatments |  |  |  |  |
| Drugs approved for PAH | 468 (93.8) | 48 (94.1) | 420 (93.8) | 0.918 |
| BPA or PEA | 273 (54.7) | 25 (49.0) | 248 (55.4) | 0.389 |
| BPA | 198 (39.7) | 18 ( 35.3) | 180 (40.2) | 0.499 |
| PEA | 106 (21.2) | 13 (25.5) | 93 (20.8) | 0.434 |

*P* values in bold represent statistical significance of Student's t-test or Mann-Whitney U-test for quantitative variables and fisher exact test for qualitative variables between CHIP and non-CHIP groups.
6MWD = 6-minute walking distance; APS = antiphospholipid syndrome; BPA = balloon pulmonary angioplasty; CHIP = clonal hematopoiesis of indeterminate potential; CI = cardiac index; CTEPH = chronic thromboembolic pulmonary hypertension; Hgb = hemoglobin; mPAP = mean pulmonary artery pressure; NT-proBNP = N-terminal pro-B-type natriuretic peptide; PAWP = pulmonary artery wedge pressure; PEA = pulmonary endarterectomy; PH = pulmonary hypertension; Plt = platelet; PVR = pulmonary vascular resistance; RAP= right atrial pressure; SvO<sub>2</sub> = mixed venous oxygen saturation; VTE = venous thromboembolism; WBC = white blood cell; WHO FC = World Health Organization functional class.

**Table 2.** Results of multivariable Cox proportional hazards models examining the relation of CHIP to the risk of all-cause death.

|  | CHIP vs Non-CHIP<br>HR (95% CI) | *P* |
| --- | --- | --- |
| Model 1 | 2.290 (1.428 - 3.672) | <0.001 |
| Model 2 | 2.161 (1.342 - 3.479) | 0.002 |
| Model 3 | 2.636 (1.415 - 4.912) | 0.002 |
| Model 4 | 3.447 (1.747 - 6.803) | <0.001 |

Model 1:unadjusted; Model 2: adjusted for age and sex; Model 3: adjusted for age, sex, 6MWD, WHO FC, NT-proBNP, RAP, SvO<sub>2</sub>, PVR and CI; Model 4: adjusted for age, sex, 6MWD, WHO FC, NT-proBNP, RAP, SvO<sub>2</sub>, PVR, CI and treatment regimens. Treatment regimens represents whether or not patients underwent PEA/BPA treatments. 6MWD = 6-minute walking distance; CHIP = clonal hematopoiesis of indeterminate potential; CI = cardiac index; HR = hazard ratio; NT-proBNP = N-terminal pro-B-type natriuretic peptide; PVR = pulmonary vascular resistance; RAP= right atrial pressure; SvO<sub>2</sub> = mixed venous oxygen saturation; WHO FC = World Health Organization functional class.

### Prevalence of CHIP

Fifty-one (10.2%) patients were found to carry one or more of the CHIP mutations at a VAF of ≥ 2%, with six of them carrying two mutations (Figure 2A). With an identical analysis strategy, CTEPH patients in our cohort presented a higher prevalence than the healthy population of comparative age in the “All of Us” Research Program, suggesting that CHIP mutations are enriched in patients with CTEPH.

**Figure 2.**
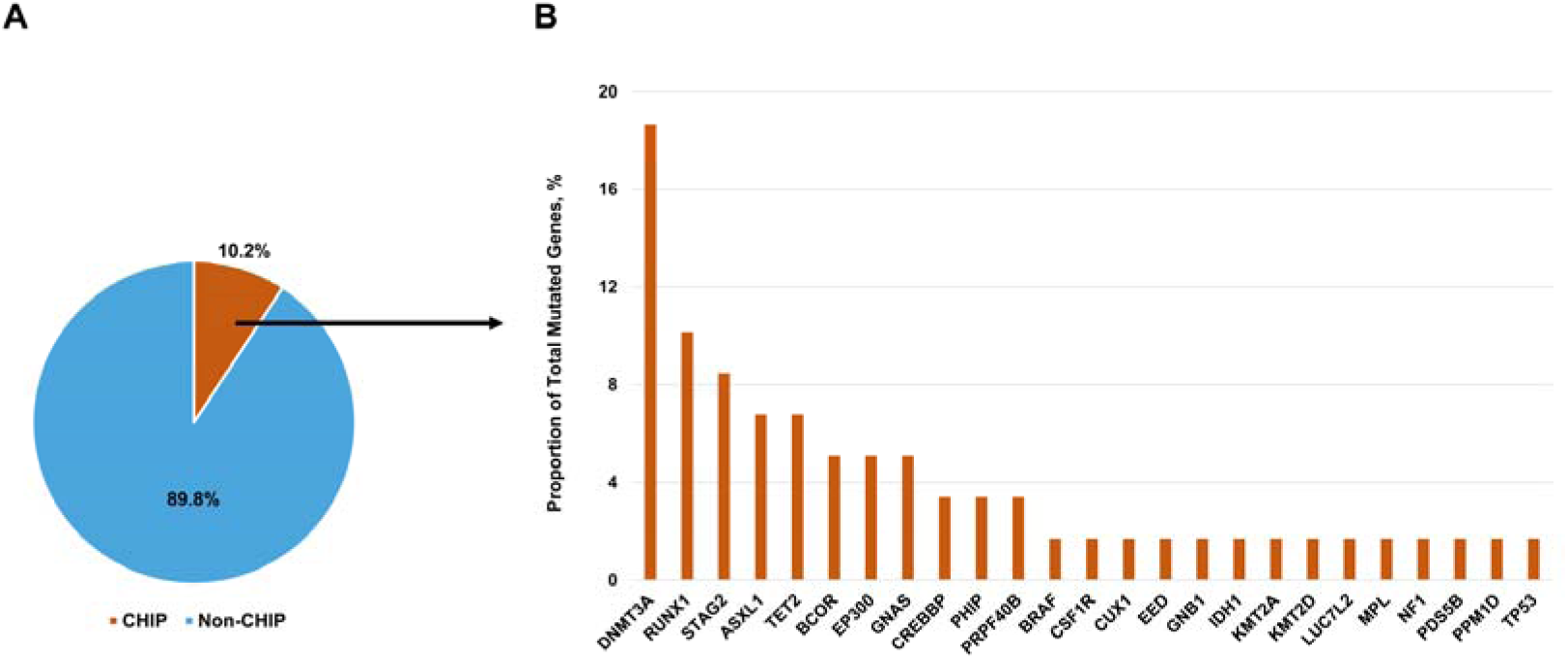
The proportion of CHIP-related mutated genes in patients with chronic thromboembolic pulmonary hypertension. CHIP = clonal hematopoiesis of indeterminate potential.

A total of 52 mutations in 25 genes were identified, and the somatic mutations most commonly occurred in the genes *DNMT3A* (11 patients), *RUNX1* (six patients), *STAG2* (five patients), followed by *TET2*, *ASXL1*, *BCOR*, *EP300* and *GNAS* (three patients each). Mutations in *CREBBP, PHIP* and *PRPF40B* were identified in two patients, and 14 other genes associated with CHIP were found to be mutated in individual patients (Figure 2B). A detailed list including types of mutations and VAFs is provided in the Supplemental Material, Table S2.

Compared with non-carriers, patients with CHIP mutations were older, had significantly higher N-terminal pro-B-type natriuretic peptide (NT-proBNP) and D-dimer levels, decreased cardiac index (CI), mixed venous oxygen saturation (SvO2), and 6MWD, and more compromised cardiac function. Other clinical characteristics as well as treatment regimens showed no difference between the two groups (Table 1).

### Follow-up

With 18 patients lost for follow-up, a total of 481 patients were further included in the survival analysis. During a median follow-up of 55 months, deaths occurred in 21 patients (42.9%) in the CHIP group and 105 patients (24.3%) in the non-CHIP group, contributing to the 5-year survival rate of 65.3% in the CHIP group and 81.9% in the non-CHIP group (*P* < 0.001 for the log-rank test) (Figure 3). In univariate analysis, the presence of CHIP mutations with a VAF of ≥ 2% was significantly associated with all-cause mortality with HR of 2.290 (95% CI: 1.428 – 3.672; *P* < 0.001). After adjustment for age and sex or in combination with clinical and hemodynamic characteristics, the association remained robust (HR: 2.161, 95% CI: 1.342 – 3.479, *P* = 0.002; HR: 2.636, 95% CI: 1.415 – 4.912, *P* = 0.002). Finally, by multivariate Cox proportional regression analysis, including age, sex, clinical and hemodynamic characteristics and treatment regimens (whether or not patients underwent PEA/BPA treatments) as covariants, all-cause death still remained independently associated with the presence of somatic mutations associated with CHIP (HR: 3.447; 95% CI: 1.747 – 6.803; *P* < 0.001) (Table 3). There was no evidence of effect modification across different treatment groups (*P* for interaction = 0.886).

**Figure 3.**
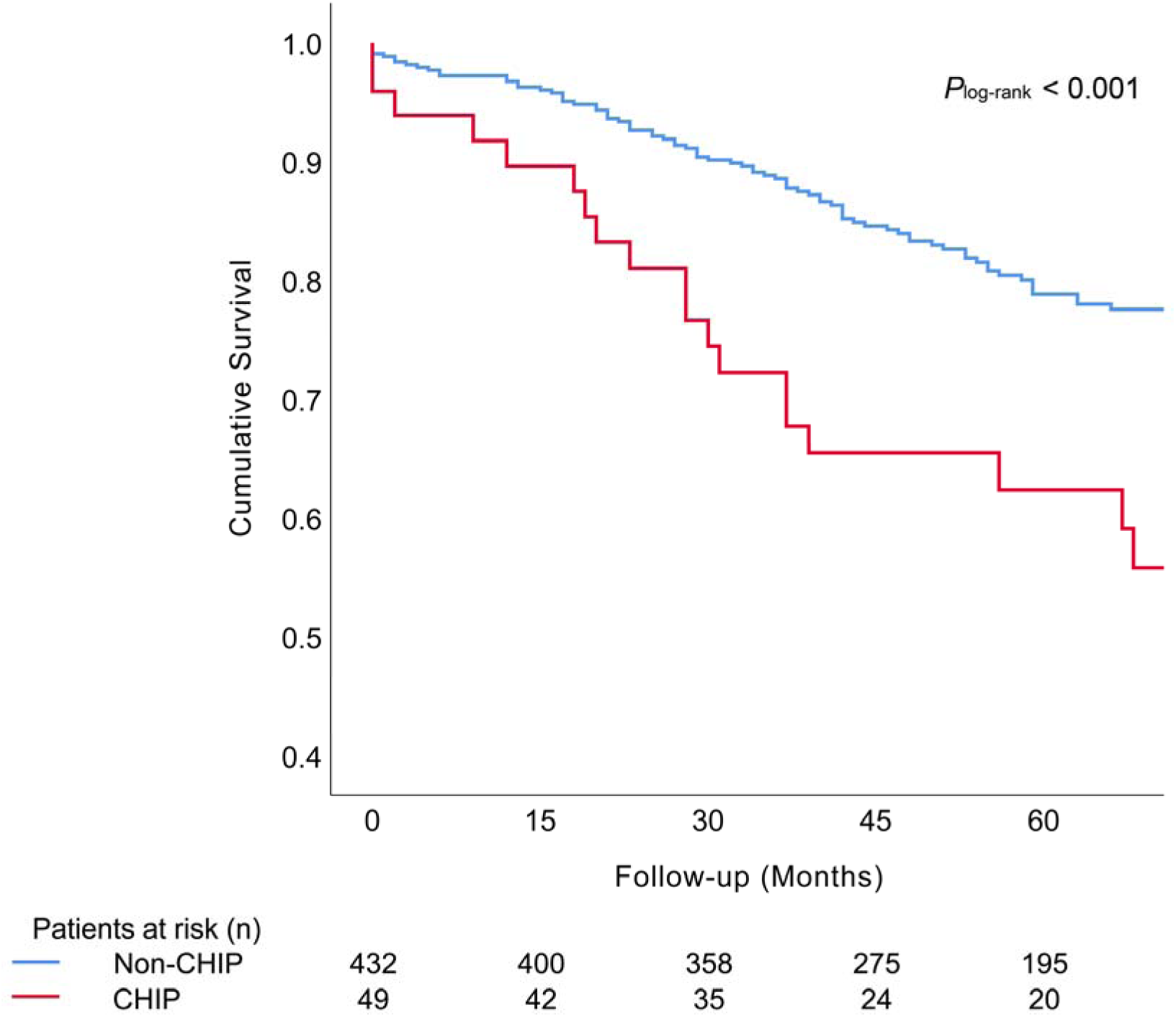
Kaplan-Meier survival curve for all-cause death stratified by CHIP status in chronic thromboembolic pulmonary hypertension. CHIP = clonal hematopoiesis of indeterminate potential.

**Figure 4.**
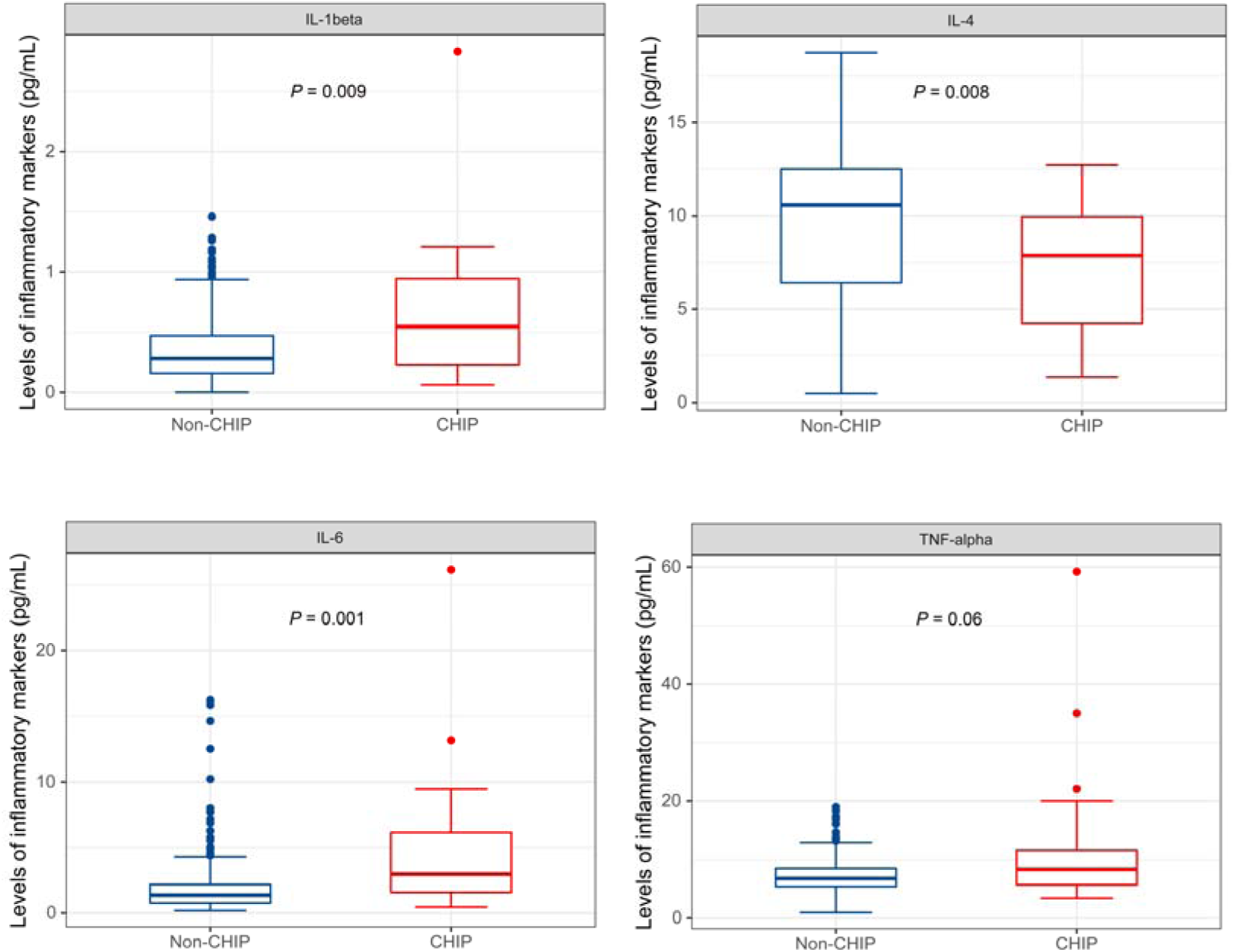
Inflammatory cytokines characteristics in patients of CHIP group/non-CHIP group. CHIP = clonal hematopoiesis of indeterminate potential.

**Figure 5.**
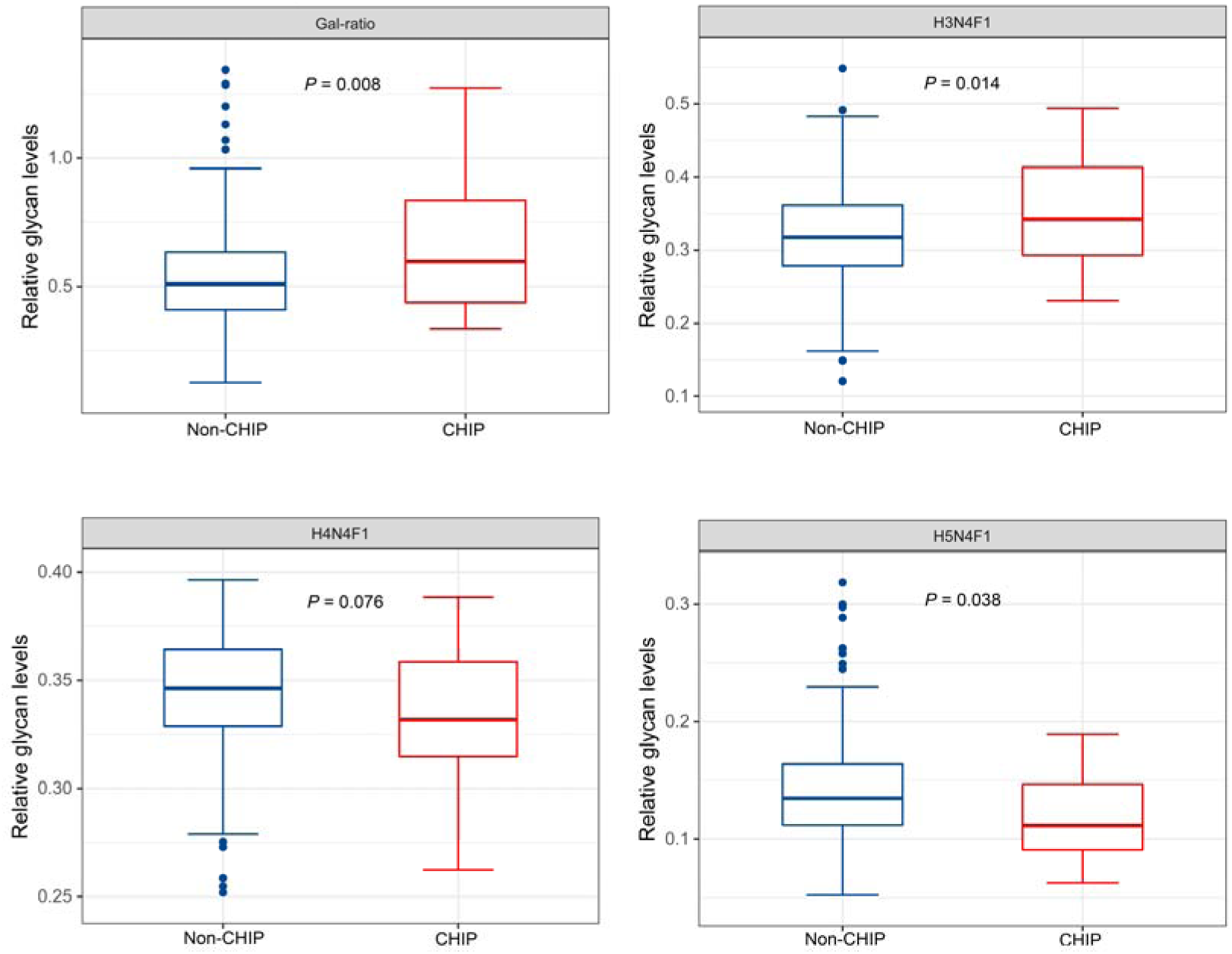
Inflammatory markers of IgG galactosylation distribution in CHIP group/non-CHIP group patients. CHIP = clonal hematopoiesis of indeterminate potential.

### Potential Mechanistic Insights

Mechanistically, the most common mutated genes in CHIP, *DNMT3A* and *TET2*, were experimentally shown to regulate the inflammatory potential of circulating leucocytes^19, 28–30^. Meanwhile, various immune cells as well as chemokines were enriched in plasma or thrombotic lesions of CTEPH patients^3–6^. In the present study, multiple inflammatory mediators including TNF-alpha, IL-1beta, IL-4 and IL-6 were measured for 20 patients carrying CHIP mutations and 201 patients without such. In line with previous works^31, 32^, our data showed elevated pro-inflammatory cytokines IL-1beta (*P* = 0.009), IL-6 (*P* = 0.001) and TNF-alpha (*P* = 0.06) and decreased anti-inflammatory cytokine IL-4 (*P* = 0.008) in the CHIP group of CTEPH patients (Figure 3).

Besides, our previous study suggested that IgG N-glycan characteristics of CTEPH patients presented an obvious pro-inflammatory phenotype (decreased proportion of IgG galactosylation [higher IgG Gal-ratio])^33^. Decreased IgG galactosylation has been reported to be a hallmark of inflammation^34^. In the present study, baseline IgG glycan characteristics were analysed in a subset of 221 patients, of whom 20 carried CHIP mutations. Glycan analysis was performed without knowing the CHIP status of the patients. We found the level of H5N4F1 (*P* = 0.038) was significantly lower and the levels of H3N4F1 (*P* = 0.014) and IgG Gal-ratio (*P* = 0.008) (Gal-ratio = H3N4F1/[H4N4F1+2*H5N4F1]) were significantly higher in the CHIP group than in the non-CHIP group.

## Discussion

The results of the present study significantly extend previous reports showing that CHIP-driver mutations are associated with the risk and prognosis of patients with cardiovascular diseases including coronary artery disease^19^, chronic heart failure^35, 36^ and degenerative aortic valve stenosis^37^. CHIP-related studies in PH were quite rare and previous studies focused only on the relationship between specific driver mutations (*TET2* and *JAK2V617F*)^10, 38, 39^ and the risk of PH. To the best of our knowledge, this is the first study focused on a well-defined group of CTEPH patients to reveal an overview of CHIP-driver genes mutation with non-biased genome sequencing and to investigate its relationship with the prognosis of CTEPH patients. Our results demonstrate that CHIP-driver mutations occurred frequently in this patient population, were associated with more severe inflammatory states, and conferred profoundly worse outcomes.

CHIP mutations can be identified in peripheral blood samples sequenced using approaches that cover the whole genome, whole exome or targeted genetic regions; however, differentiating true CHIP mutations from sequencing artifacts and germline variants is a considerable bioinformatic challenge. To increase the accuracy of CHIP calls, we referred to a step-wise method raised by Bick *et al.*^23^. This method combines filtering based on sequencing metrics, variant annotation, and population-based associations method to increase the fidelity of CHIP calls. With an identical calling approach, the prevalence of CHIP mutations in CTEPH patients is higher than in the age-paired healthy population in the “All of Us” Research Program, suggesting that CHIP mutations are enriched in patients with CTEPH. Interestingly, the most common mutations in this population are *DNMT3A*, *RUNX1* and *STAG2*, followed by *TET2* and other mutations, presenting a bit little different scenario from most previous reports. This difference could be blamed on the discrepancy in either diseases studied, population or sequencing and calling approaches.

In this study, the association of CHIP with mortality was observed in a univariate model. Consistent with previous studies, patients in the CHIP group are significantly older than the non-CHIP group, and CHIP effects were robust after adjustment for age and sex. NT-proBNP, CI, SvO_2_, WHO FC and 6MWD showed significant differences between the CHIP and non-CHIP groups, which may simply be a reflection of the age difference between the groups. WHO FC, 6MWD, NT-proBNP, SvO_2_, right atrial pressure, CI, pulmonary vascular resistance and treatment regimens as well as age and sex were used to further adjust the model. Importantly, all-cause death still remained independently associated with the presence of somatic mutations associated with CHIP.

Recent studies based on CANTOS (Canakinumab Anti-Inflammatory Thrombosis Outcome Study) and UK Biobank reported that patients harboring *TET2* mutations showed a statistically significant reduced risk for incident major cardiovascular events by canakinumab, indicating that some patients with CHIP mutations especially *TET2* could potentially benefit from targeted anti-inflammatory therapy^40, 41^. This is in line with previous works demonstrating increased levels of IL-6 and IL-1beta in *Tet2* loss-of-function mouse models^19, 28^. In this study, levels of IL-1beta and IL-6 are higher in CHIP patients, implicating targeted anti-inflammatory therapy might benefit the subgroup of CTEPH patients with CHIP mutations.

Aside from inflammation mediated by myeloid cells, lymphoid cell-related immune activation was also found to be involved in CHIP-related pro-inflammatory effects. Mas-Peiro *et al.* reported that Th17 to regulatory T-cell ratios were augmented in aortic stenosis patients with *DNMT3A* mutations^37^. What’s more, circulating monocytes and T cells of patients with heart failure harboring clonal hematopoiesis-driver mutations in *DNMT3A* exhibit a highly inflamed transcriptome^42^.

These may be in line with different lineage restrictions, which indicate a myeloid bias for *TET2*, but multipotent stem cell origin for *DNMT3A*^43^. *DNMT3A*-driven mutations prevailed in the patients of our cohort, so we further test the pro-inflammatory effects of CHIP from lymphoid perspectives.

IgG, the most abundant glycoprotein secreted by lymphoid cells and the main effector of humoral immunity^44^, plays a central role in human immune responses and is involved in inflammatory pathways. IgG regulates systemic inflammatory homeostasis at multiple levels primarily by its N-glycans^45^. Decreased galactosylation of IgG N-glycans is reported as a hallmark of inflammation^34^. Moreover, our preliminary research has found that IgG N-glycome in CTEPH presented a pro-inflammatory phenotype (up-regulated IgG Gal-ratio representing decreased IgG galactosylation), compared with healthy controls and IgG Gal-ratio is positively correlated with the level of NT-proBNP^33^. In this study, we found that patients in the CHIP group showed a more severe inflammatory state reflected by up-regulated IgG Gal-ratio, compared with that in the non-CHIP group, suggesting a probable relationship between CHIP and secreted IgG antibody phenotype. However, the causality and directionality of the link between the pro-inflammatory function of IgG and CHIP still require further investigation.

## Limitations

Firstly, the current study focused on a thoroughly annotated clinical cohort of patients with CTEPH. Although this is a comparably large clinical cohort of patients with CTEPH, the sample size is still limited. Secondly, considering the sensitivity of next-generation sequencing, the presence of CHIP was defined as the occurrence of mutations with a VAF of at least 0.02 in this study. However, the results of previous studies indicate that even lower VAFs may be associated with the development of various cardiovascular diseases. Further studies with more sensitive sequencing approaches are required to identify more specific thresholds of VAF for CTEPH patients. Lastly, the involvement of CHIP in the pathogenesis of CTEPH still needs to be further clarified in the future.

## Conclusion

Our data demonstrated that CHIP mutations were enriched in CTEPH patients and represented an additional disease component in this disease that independently impacts prognosis.

## Data Availability

The data supporting the results of the paper is provided in the manuscript. Other data underlying this article will be shared on reasonable requests to the corresponding authors.

## Non-standard Abbreviations and Acronyms

6MWD: 6-minute walking distance
BPA: balloon pulmonary angioplasty
CHIP: clonal hematopoiesis of indeterminate potential
CI: cardiac index
CTEPH: chronic thromboembolic pulmonary hypertension
MALDI-TOF-MS: matrix-assisted laser desorption/ionization time of flight mass spectrometry
MPN: myeloproliferative neoplasms
NT-proBNP: N-terminal pro-B-type natriuretic peptide
PEA: pulmonary endarterectomy
PH: pulmonary hypertension
SvO_2_: mixed venous oxygen saturation
VAF: variant allele frequency
WES: whole-exome sequencing
WGS: whole-genome sequencing
WHO FC: World Health Organization functional class

## Acknowledgements

Author Contributions

Conception and design: ZCJ, ZJZ, CL, TYL, CYC; Analysis and interpretation of data: CL, ZJZ, LHQ, BCQ, LYR, JSM; Drafting of the manuscript: CL, ZJZ, YJY, CYC, YPW, YJZ; critical revision of the manuscript for important intellectual content: ZCJ, YPZ, TYL, SJZ; final approval of the manuscript: all authors; statistical expertise: DLW, KS; obtaining of research funding: ZCJ, ZJZ, YPZ; acquisition of data: ZYH, JHL, QHZ, LW, XQX, RNL, JW.

## Supplemental Materials

Tables S1-S2

## References

1. Delcroix M, Torbicki A, Gopalan D, Sitbon O, Klok FA, Lang I, Jenkins D, Kim NH, Humbert M, Jais X, et al. ERS statement on chronic thromboembolic pulmonary hypertension. Eur Respir J. 2021;57. doi: 10.1183/13993003.02828-2020

2. Simonneau G, Montani D, Celermajer DS, Denton CP, Gatzoulis MA, Krowka M, Williams PG, Souza R. Haemodynamic definitions and updated clinical classification of pulmonary hypertension. Eur Respir J. 2019;53. doi: 10.1183/13993003.01913-2018

3. Quarck R, Wynants M, Verbeken E, Meyns B, Delcroix M. Contribution of inflammation and impaired angiogenesis to the pathobiology of chronic thromboembolic pulmonary hypertension. Eur Respir J. 2015;46:431–443. doi: 10.1183/09031936.00009914

4. Bochenek ML, Rosinus NS, Lankeit M, Hobohm L, Bremmer F, Schutz E, Klok FA, Horke S, Wiedenroth CB, Munzel T, et al. From thrombosis to fibrosis in chronic thromboembolic pulmonary hypertension. Thromb Haemost. 2017;117:769–783. doi: 10.1160/TH16-10-0790

5. Zabini D, Heinemann A, Foris V, Nagaraj C, Nierlich P, Balint Z, Kwapiszewska G, Lang IM, Klepetko W, Olschewski H, et al. Comprehensive analysis of inflammatory markers in chronic thromboembolic pulmonary hypertension patients. Eur Respir J. 2014;44:951–962. doi: 10.1183/09031936.00145013

6. Yang M, Deng C, Wu D, Zhong Z, Lv X, Huang Z, Lian N, Liu K, Zhang Q. The role of mononuclear cell tissue factor and inflammatory cytokines in patients with chronic thromboembolic pulmonary hypertension. J Thromb Thrombolysis. 2016;42:38–45. doi: 10.1007/s11239-015-1323-2

7. Zia A, Russell J, Sarode R, Veeram SR, Josephs S, Malone K, Zhang S, Journeycake J. Markers of coagulation activation, inflammation and fibrinolysis as predictors of poor outcomes after pediatric venous thromboembolism: A systematic review and meta-analysis. Thromb Res. 2017;160:1–8. doi: 10.1016/j.thromres.2017.10.003

8. Satoh T, Satoh K, Yaoita N, Kikuchi N, Omura J, Kurosawa R, Numano K, Al-Mamun E, Siddique MA, Sunamura S, et al. Activated TAFI Promotes the Development of Chronic Thromboembolic Pulmonary Hypertension: A Possible Novel Therapeutic Target. Circ Res. 2017;120:1246–1262. doi: 10.1161/CIRCRESAHA.117.310640

9. Yaoita N, Satoh K, Satoh T, Shimizu T, Saito S, Sugimura K, Tatebe S, Yamamoto S, Aoki T, Kikuchi N, et al. Identification of the Novel Variants in Patients With Chronic Thromboembolic Pulmonary Hypertension. J Am Heart Assoc. 2020;9:e015902. doi: 10.1161/JAHA.120.015902

10. Eichstaedt CA, Verweyen J, Halank M, Benjamin N, Fischer C, Mayer E, Guth S, Wiedenroth CB, Egenlauf B, Harutyunova S, et al. Myeloproliferative Diseases as Possible Risk Factor for Development of Chronic Thromboembolic Pulmonary Hypertension-A Genetic Study. Int J Mol Sci. 2020;21. doi: 10.3390/ijms21093339

11. Hasselbalch HC, Elvers M, Schafer AI. The pathobiology of thrombosis, microvascular disease, and hemorrhage in the myeloproliferative neoplasms. Blood. 2021;137:2152–2160. doi: 10.1182/blood.2020008109

12. De Stefano V, Ruggeri M, Cervantes F, Alvarez-Larran A, Iurlo A, Randi ML, Elli E, Finazzi MC, Finazzi G, Zetterberg E, et al. High rate of recurrent venous thromboembolism in patients with myeloproliferative neoplasms and effect of prophylaxis with vitamin K antagonists. Leukemia. 2016;30:2032–2038. doi: 10.1038/leu.2016.85

13. Venton G, Turcanu M, Colle J, Thuny F, Chebrek S, Farnault L, Mercier C, Ivanov V, Fanciullino R, Suchon P, et al. Pulmonary hypertension in patients with myeloproliferative neoplasms: A large cohort of 183 patients. Eur J Intern Med. 2019;68:71–75. doi: 10.1016/j.ejim.2019.08.004

14. Genty T, Wirth C, Humbert M, Fadel E, Stephan F. Pulmonary Endarterectomy in Patients With Myeloproliferative Neoplasms. Chest. 2022;161:552–556. doi: 10.1016/j.chest.2021.09.007

15. Steensma DP, Bejar R, Jaiswal S, Lindsley RC, Sekeres MA, Hasserjian RP, Ebert BL. Clonal hematopoiesis of indeterminate potential and its distinction from myelodysplastic syndromes. Blood. 2015;126:9–16. doi: 10.1182/blood-2015-03-631747

16. Leiva O, Hobbs G, Ravid K, Libby P. Cardiovascular Disease in Myeloproliferative Neoplasms: JACC: CardioOncology State-of-the-Art Review. JACC CardioOncol. 2022;4:166–182. doi: 10.1016/j.jaccao.2022.04.002

17. Jaiswal S, Ebert BL. Clonal hematopoiesis in human aging and disease. Science. 2019;366. doi: 10.1126/science.aan4673

18. Jaiswal S, Fontanillas P, Flannick J, Manning A, Grauman PV, Mar BG, Lindsley RC, Mermel CH, Burtt N, Chavez A, et al. Age-related clonal hematopoiesis associated with adverse outcomes. N Engl J Med. 2014;371:2488–2498. doi: 10.1056/NEJMoa1408617

19. Jaiswal S, Natarajan P, Silver AJ, Gibson CJ, Bick AG, Shvartz E, McConkey M, Gupta N, Gabriel S, Ardissino D, et al. Clonal Hematopoiesis and Risk of Atherosclerotic Cardiovascular Disease. N Engl J Med. 2017;377:111–121. doi: 10.1056/NEJMoa1701719

20. Honigberg MC, Zekavat SM, Niroula A, Griffin GK, Bick AG, Pirruccello JP, Nakao T, Whitsel EA, Farland LV, Laurie C, et al. Premature Menopause, Clonal Hematopoiesis, and Coronary Artery Disease in Postmenopausal Women. Circulation. 2021;143:410–423. doi: 10.1161/CIRCULATIONAHA.120.051775

21. Yu B, Roberts MB, Raffield LM, Zekavat SM, Nguyen NQH, Biggs ML, Brown MR, Griffin G, Desai P, Correa A, et al. Supplemental Association of Clonal Hematopoiesis With Incident Heart Failure. J Am Coll Cardiol. 2021;78:42–52. doi: 10.1016/j.jacc.2021.04.085

22. Galie N, Humbert M, Vachiery JL, Gibbs S, Lang I, Torbicki A, Simonneau G, Peacock A, Vonk Noordegraaf A, Beghetti M, et al. 2015 ESC/ERS Guidelines for the diagnosis and treatment of pulmonary hypertension: The Joint Task Force for the Diagnosis and Treatment of Pulmonary Hypertension of the European Society of Cardiology (ESC) and the European Respiratory Society (ERS): Endorsed by: Association for European Paediatric and Congenital Cardiology (AEPC), International Society for Heart and Lung Transplantation (ISHLT). Eur Heart J. 2016;37:67–119. doi: 10.1093/eurheartj/ehv317

23. Vlasschaert C, Mack T, Heimlich JB, Niroula A, Uddin MM, Weinstock J, Sharber B, Silver AJ, Xu Y, Savona M, et al. A practical approach to curate clonal hematopoiesis of indeterminate potential in human genetic data sets. Blood. 2023;141:2214–2223. doi: 10.1182/blood.2022018825

24. Cibulskis K, Lawrence MS, Carter SL, Sivachenko A, Jaffe D, Sougnez C, Gabriel S, Meyerson M, Lander ES, Getz G. Sensitive detection of somatic point mutations in impure and heterogeneous cancer samples. Nat Biotechnol. 2013;31:213–219. doi: 10.1038/nbt.2514

25. Tang J, Liu H, Sun M, Zhang X, Chu H, Li Q, Prokosch V, Cui H. Aqueous Humor Cytokine Response in the Contralateral Eye after First-Eye Cataract Surgery in Patients with Primary Angle-Closure Glaucoma, High Myopia or Type 2 Diabetes Mellitus. Front Biosci (Landmark Ed). 2022;27:222. doi: 10.31083/j.fbl2707222

26. Ren S, Zhang Z, Xu C, Guo L, Lu R, Sun Y, Guo J, Qin R, Qin W, Gu J. Distribution of IgG galactosylation as a promising biomarker for cancer screening in multiple cancer types. Cell Res. 2016;26:963–966. doi: 10.1038/cr.2016.83

27. Qian Y, Wang Y, Zhang X, Zhou L, Zhang Z, Xu J, Ruan Y, Ren S, Xu C, Gu J. Quantitative analysis of serum IgG galactosylation assists differential diagnosis of ovarian cancer. J Proteome Res. 2013;12:4046–4055. doi: 10.1021/pr4003992

28. Fuster JJ, MacLauchlan S, Zuriaga MA, Polackal MN, Ostriker AC, Chakraborty R, Wu CL, Sano S, Muralidharan S, Rius C, et al. Clonal hematopoiesis associated with TET2 deficiency accelerates atherosclerosis development in mice. Science. 2017;355:842–847. doi: 10.1126/science.aag1381

29. Sano S, Oshima K, Wang Y, Katanasaka Y, Sano M, Walsh K. CRISPR-Mediated Gene Editing to Assess the Roles of Tet2 and Dnmt3a in Clonal Hematopoiesis and Cardiovascular Disease. Circ Res. 2018;123:335–341. doi: 10.1161/CIRCRESAHA.118.313225

30. Sano S, Oshima K, Wang Y, MacLauchlan S, Katanasaka Y, Sano M, Zuriaga MA, Yoshiyama M, Goukassian D, Cooper MA, et al. Tet2-Mediated Clonal Hematopoiesis Accelerates Heart Failure Through a Mechanism Involving the IL-1beta/NLRP3 Inflammasome. J Am Coll Cardiol. 2018;71:875–886. doi: 10.1016/j.jacc.2017.12.037

31. Bohme M, Desch S, Rosolowski M, Scholz M, Krohn K, Buttner P, Cross M, Kirchberg J, Rommel KP, Poss J, et al. Impact of Clonal Hematopoiesis in Patients With Cardiogenic Shock Complicating Acute Myocardial Infarction. J Am Coll Cardiol. 2022;80:1545–1556. doi: 10.1016/j.jacc.2022.08.740

32. Abplanalp WT, Mas-Peiro S, Cremer S, John D, Dimmeler S, Zeiher AM. Association of Clonal Hematopoiesis of Indeterminate Potential With Inflammatory Gene Expression in Patients With Severe Degenerative Aortic Valve Stenosis or Chronic Postischemic Heart Failure. JAMA Cardiol. 2020;5:1170–1175. doi: 10.1001/jamacardio.2020.2468

33. Zhang Z, Wang H, Lian T, Zhou Y, Xu X, Guo F, Wei Y, Li J, Sun K, Liu C, et al. Human Plasma Immunoglobulin G N-Glycome Profiles Reveal a Pro-Inflammatory Phenotype in Chronic Thromboembolic Pulmonary Hypertension: A Cross-Sectional Study of Two Independent Cohorts. SSRN preprint. 2023. doi: 10.2139/ssrn.4366881

34. Plomp R, Ruhaak LR, Uh HW, Reiding KR, Selman M, Houwing-Duistermaat JJ, Slagboom PE, Beekman M, Wuhrer M. Subclass-specific IgG glycosylation is associated with markers of inflammation and metabolic health. Sci Rep. 2017;7:12325. doi: 10.1038/s41598-017-12495-0

35. Dorsheimer L, Assmus B, Rasper T, Ortmann CA, Ecke A, Abou-El-Ardat K, Schmid T, Brune B, Wagner S, Serve H, et al. Association of Mutations Contributing to Clonal Hematopoiesis With Prognosis in Chronic Ischemic Heart Failure. JAMA Cardiol. 2019;4:25–33. doi: 10.1001/jamacardio.2018.3965

36. Pascual-Figal DA, Bayes-Genis A, Diez-Diez M, Hernandez-Vicente A, Vazquez-Andres D, de la Barrera J, Vazquez E, Quintas A, Zuriaga MA, Asensio-Lopez MC, et al. Clonal Hematopoiesis and Risk of Progression of Heart Failure With Reduced Left Ventricular Ejection Fraction. J Am Coll Cardiol. 2021;77:1747–1759. doi: 10.1016/j.jacc.2021.02.028

37. Mas-Peiro S, Hoffmann J, Fichtlscherer S, Dorsheimer L, Rieger MA, Dimmeler S, Vasa-Nicotera M, Zeiher AM. Clonal haematopoiesis in patients with degenerative aortic valve stenosis undergoing transcatheter aortic valve implantation. Eur Heart J. 2020;41:933–939. doi: 10.1093/eurheartj/ehz591

38. Potus F, Pauciulo MW, Cook EK, Zhu N, Hsieh A, Welch CL, Shen Y, Tian L, Lima P, Mewburn J, et al. Novel Mutations and Decreased Expression of the Epigenetic Regulator TET2 in Pulmonary Arterial Hypertension. Circulation. 2020;141:1986–2000. doi: 10.1161/CIRCULATIONAHA.119.044320

39. Kimishima Y, Misaka T, Yokokawa T, Wada K, Ueda K, Sugimoto K, Minakawa K, Nakazato K, Ishida T, Oshima M, et al. Clonal hematopoiesis with JAK2V617F promotes pulmonary hypertension with ALK1 upregulation in lung neutrophils. Nat Commun. 2021;12:6177. doi: 10.1038/s41467-021-26435-0

40. Svensson EC, Madar A, Campbell CD, He Y, Sultan M, Healey ML, Xu H, D’Aco K, Fernandez A, Wache-Mainier C, et al. TET2-Driven Clonal Hematopoiesis and Response to Canakinumab: An Exploratory Analysis of the CANTOS Randomized Clinical Trial. JAMA Cardiol. 2022;7:521–528. doi: 10.1001/jamacardio.2022.0386

41. Vlasschaert C, Heimlich JB, Rauh MJ, Natarajan P, Bick AG. Interleukin-6 Receptor Polymorphism Attenuates Clonal Hematopoiesis-Mediated Coronary Artery Disease Risk Among 451 180 Individuals in the UK Biobank. Circulation. 2023;147:358–360. doi: 10.1161/CIRCULATIONAHA.122.062126

42. Abplanalp WT, Cremer S, John D, Hoffmann J, Schuhmacher B, Merten M, Rieger MA, Vasa-Nicotera M, Zeiher AM, Dimmeler S. Clonal Hematopoiesis-Driver DNMT3A Mutations Alter Immune Cells in Heart Failure. Circ Res. 2021;128:216–228. doi: 10.1161/CIRCRESAHA.120.317104

43. Buscarlet M, Provost S, Zada YF, Bourgoin V, Mollica L, Dube MP, Busque L. Lineage restriction analyses in CHIP indicate myeloid bias for TET2 and multipotent stem cell origin for DNMT3A. Blood. 2018;132:277–280. doi: 10.1182/blood-2018-01-829937

44. Vidarsson G, Dekkers G, Rispens T. IgG subclasses and allotypes: from structure to effector functions. Front Immunol. 2014;5:520. doi: 10.3389/fimmu.2014.00520

45. Gudelj I, Lauc G, Pezer M. Immunoglobulin G glycosylation in aging and diseases. Cell Immunol. 2018;333:65–79. doi: 10.1016/j.cellimm.2018.07.009

